# Chinese Herbal Medicine as a complementary therapy for the management of Colorectal Cancer: Study protocol for a Delphi Expert Consensus survey

**DOI:** 10.64898/2026.04.21.26350990

**Authors:** Chester Yan Jie Ng, Ming Liu, Dandan Ai, Liang Yao, Mingxiao Yang, Linda LD Zhong

**Affiliations:** School of Biological Sciences, Nanyang Technological University, 60 Nanyang Drive, Singapore 637551; Lee Kong Chian School of Medicine, Nanyang Technological University, Experimental Medicine Building, 59 Nanyang Drive, Singapore, 636921

**Keywords:** Chinese Medicine, Chinese Herbal Medicine, Colorectal Cancer, Consensus, Protocol, Delphi

## Abstract

**Introduction:** Colorectal cancer (CRC) remains a leading cause of cancer-related morbidity and mortality worldwide, despite advances in conventional oncological therapies. In recent years, various studies have made advances in integrative oncology, such as investigating the use of Chinese Herbal Medicine (CHM) as a complementary therapy alongside conventional oncological therapies to alleviate treatment-related adverse effects, improve quality of life, and potentially enhance therapeutic outcomes. Despite this, clinical practice in this area remains highly heterogeneous, with limited standardized guidelines on key areas of concern such as (1) optimal intervention, (2) recommended stage and duration of intervention, (3) safety considerations, and (4) possible herb-drug interactions. Hence, this study aims to establish expert consensus on the usage of CHM as a complementary therapy in the management of CRC, to support safe, consistent, and evidence-informed clinical practice.

**Methods and Analysis:** We will employ a modified Delphi technique to achieve consensus amongst a panel of international experts in various fields related to integrative oncology. Prior to the study, a list of questionnaire items was developed based on a systematic review of existing clinical practice guidelines on CRC. An international panel will be invited based on established international profile in integrative oncology research and clinical practice, and by peer referral. Two rounds of Delphi will be conducted using anonymous online questionnaires. Consensus will be considered reached if at least 50% of the panel strongly agree/disagree that an item should be included or excluded while strong consensus will be set at 76%. Items which achieve strong consensus after Round 1 will be removed, before being sent out for Round 2 with a summary of Round 1 responses for a final consensus.

**Ethics and Dissemination:** Ethics approval has been obtained from the Institutional Review Board of Nanyang Technological University (IRB-2025-1222). Our findings will be disseminated through peer-reviewed publications and conference presentations.

**Strengths and limitations of this study:**

- This study will develop an expert consensus which aims to guide future integration of Chinese Herbal Medicine (CHM) as a complementary therapy into colorectal cancer (CRC) management.
- Key concerns in areas such as determining the (1) optimal intervention, (2) recommended stage and duration of intervention, (3) safety considerations, and (4) possible herb-drug interactions, thereby laying the groundwork for potential future incorporation of CHM into CRC treatment protocols alongside conventional oncology approaches has been identified, thus limiting implementation in clinical practice.
- Designing a study e-guide, followed by the consensus rounds study online will facilitate participants’ responses and the dissemination of information from previous rounds.

## Introduction

According to recent worldwide cancer statistics, CRC is the third most prevalent cancer in both men and women, accounting for 9.6% of total cancer diagnosis [1]. CRC also accounted for 13% of all malignant tumors, and some common clinical symptoms include abdominal pain, alteration of chronic bowel habits, changes in bowel movements, unintentional weight loss, anorexia, abdominal distension, and gastrointestinal bleeding [2-5]. Currently, the main treatment methods for CRC include surgical resection, chemotherapy, immunotherapy, and radiotherapy [6, 7]. Amongst them, chemotherapy is primarily used for patients with advanced CRC, especially in the metastatic stage [8]. However, despite the well-acknowledged benefits of chemotherapy, side-effects might arise in some patients, which can vary from nausea, vomiting, and diarrhoea to symptoms such as neuropathy, cardiomyopathy, and lung fibrosis [9-12]. Therefore, there lies a growing need to source for treatment approaches which could complement existing oncological treatments, while potentially alleviating the severity of these side-effects, to improve the quality of life (QOL) of patients with CRC.

In recent years, integrative oncology, defined by the complementary use of lifestyle modifications, mind and body therapies, and natural products to improve symptom management and quality of life among patients with cancer, have been extensively researched on [13, 14]. Amongst the various integrative oncological approaches, one potential form of complementary therapy is Chinese Medicine (CM), which has been extensively researched and has shown potential benefits in cancer management. A key treatment modality of CM is Chinese Herbal Medicine (CHM), which has recently been discovered to not only possess anticancer properties, but may also mitigate toxicity-related adverse effects caused by chemotherapeutics. For instance, studies have shown that bioactive components in CHM possess anti-cancer effects such as disruption of the tumour microenvironment, promoting cancer cell apoptosis and enhancing immunity [15, 16]. In addition, studies have also found that combining CHM with other chemotherapy agents could potentially reduce adverse reactions caused by chemotherapy, significantly improving the QOL of patients [17]. Hence, as a complementary therapy, CHM may support the management of CRC by improving QOL of patients alongside its anticancer effects [18, 19].

Despite the known benefits of how CHM could benefit the management of CRC, some concerns remain with regards to its integration in clinical practice. Key clinical concerns include determining the (1) optimal intervention, (2) recommended stage and duration of intervention, (3) safety considerations, and (4) possible herb-drug interactions, thereby laying the groundwork for potential future incorporation of CHM into CRC treatment protocols alongside conventional oncology approaches. Hence, our present study aimed to assess the feasibility of integrating CHM as a complementary therapy for CRC management via a Delphi Expert Consensus survey to establish consensus on the key clinical concerns mentioned above. We chose the Delphi technique as it is generally more flexible and adaptable to emerging methodologies, facilitate consensus among experts, and are particularly valuable when high-quality evidence is lacking [20]. This is in line with the current stage of research of CHM use in managing CRC, where high-quality evidence is still limited.

Therefore, by aiming to establish expert consensus across these domains of interest in clinical practice, this study aims to provide a structured foundation for rational and evidence-informed integration of CHM into CRC management. In addition, we also hope that the conduct of the expert consensus will allow us to identify key gaps to guide future clinical trials and translational research in CHM use for CRC management.

## Methods and Design

This study was reported in accordance with the ACCORD (ACcurate COnsensus Reporting Document), Recommendations for the Conducting and REporting of DElphi Studies (CREDES), and Delphi studies in social and health sciences—Recommendations for an interdisciplinary standardized reporting (DELPHISTAR) guidelines [21-23]. A Delphi method will be used for this study, which is a practical and structured group facilitation technique that aims to achieve consensus on the opinions of a panel of “experts” regarding an issue of concern [24]. The respondents will be tasked to participate in sequential questionnaires that constitute different rounds and each round is refined based on feedback from the previous version. After each round, group responses will be returned to the panelists, who can then reconsider their views based on this report of the group’s views. One key benefit of the Delphi method prevents situations in which the group is dominated by the views of a few [25]. This study has obtained the relevant ethics approval by the Institutional Review Board (IRB) of Nanyang Technological University (IRB-2025-1222). This protocol is not yet registered.

For this study, we intend to use a modified Delphi method consisting of 2 rounds of Delphi survey. We will use both structured statements from literature and open-ended questions in the first round in a classical Delphi method. This modified Delphi study will comprise of two stages: questionnaire development and Delphi survey. Study robustness will be enhanced by following the guideline for Conducting and REporting DELphi studies (CREDES) [22]. Prior to the start of the study, an online briefing will be recorded to brief survey participants on the conduct of the study, and the link will be sent to the survey participants to watch at their convenience. They will be briefed on the design of the study, the documents they will receive, and detailed instructions on how to fill in the survey questionnaire. Upon completion of Round 1, a written summary will be provided to the panel and using answers from Round 1, Round 2 questions (statements only) will then be re-generated and sent to the expert panel again. The proposed study flow is shown in **Figure 1** below.

**Figure 1:**
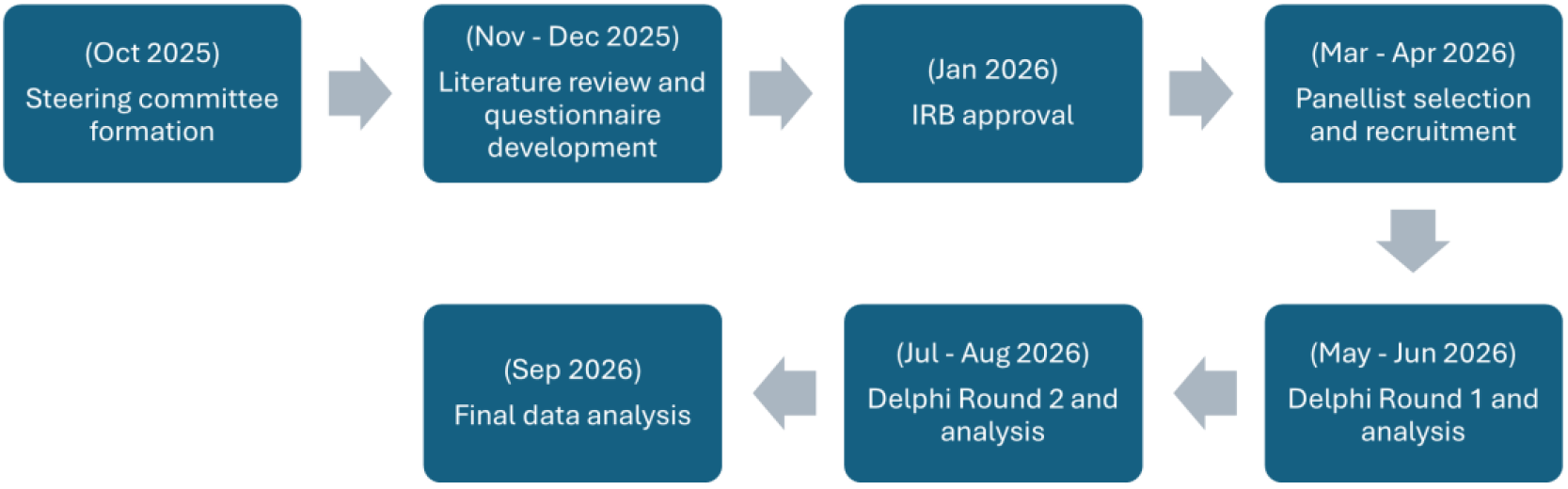
Proposed study flow

### Selection of steering committee and panellists

To guide the smooth conduct of the Expert Consensus study, a steering committee was set up (LD, MX, LY, CN, ML, DA). The steering committee consisted of experts from a variety of disciplines (epidemiology, oncology, gastroenterology, CM clinical practice) and research expertise (qualitative, quantitative, Delphi methods, guideline development). The present study methodology was established during in-person meetings and email correspondence. Agreement was reached by the steering committee regarding all key steps of study development, which included: development of the inclusion and exclusion criteria, search strategy for literature review, development of survey items, determination of consensus levels, translation of the original template items into an online survey format, reviews of survey draft iterations until final consensus of item inclusion, and statistical analysis of results. The details of our core study team are shown in **Textbox 1** below. No outside consulting in regard to method took place.

**[Textbox 1:** Core study team members]

| Steering Committee Members (Core Study Team) |
| --- |
| Chair: Assoc Prof Linda LD Zhong (LD) |
| Co-Chairs: Asst Prof Mingxiao Yang (MX), Asst Prof Liang Yao (LY) |
| Student Investigator: Chester YJ Ng (CN) |
| Study Team Members: Ming Liu (ML), Dandan Ai (DA) |

Selection of panellists for the Delphi study will be advised by the steering committee (LD, MX). Experts will be recruited globally based on their research and/or clinical practice in integrative oncology and CRC management. Based on previous recommendations, the panel size should consist of a minimum of 20 members [26-29]. The expert panellists we intend to recruit for the study will include experts in CM integrative oncology, with experience in related fields such as pharmacology research, clinical practice and policymaking. The inclusion and exclusion criteria were developed by content experts on the steering committee to ensure that selected panellists demonstrated experience through publications and/or years of clinical practice [30, 31]. A table showing the inclusion and exclusion criteria of panellists is shown in **Table 1** below. Panellists will be first identified by LD, MX and CN based on the inclusion criteria, before being invited to participate via official email correspondence. In addition, invited panellists were also asked and allowed to suggest other suitable members to join the panel, subject to final approval from the steering committee. All the experts who met the definition were invited to the first round. All the experts who completed the previous round were invited to participate in the subsequent round.

**Table 1:**
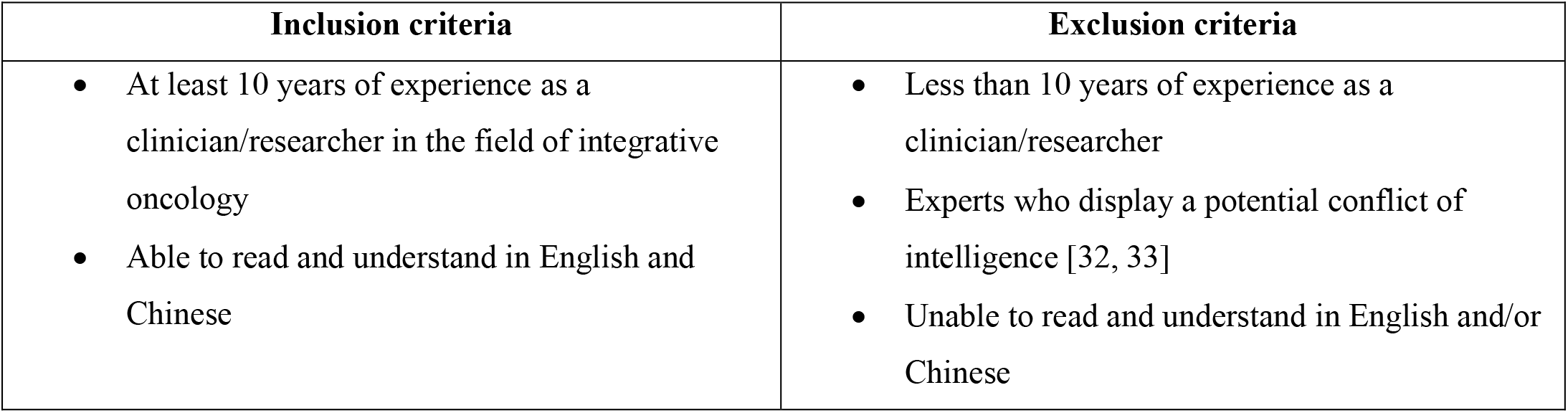
Inclusion criteria of study panellists.

| Inclusion criteria | Exclusion criteria |
| --- | --- |
| At least 10 years of experience as a clinician/researcher in the field of integrative oncology<br>Able to read and understand in English and Chinese | Less than 10 years of experience as a clinician/researcher<br>Experts who display a potential conflict of intelligence [32, 33]<br>Unable to read and understand in English and/or Chinese |

### Preparatory research / Questionnaire development

Between Sep-Nov 2025, the study team conducted three in-person meetings to discuss the optimal management methods of CRC by complementary CHM. All study team members were present at all three meetings. Patients or the public were not involved in the design, or conduct, or reporting, or dissemination plans of our research. The meetings were led by the Chair (LD) and two Co-Chairs (MX, LY) and followed the same agenda. After the first meeting, based on the discussions and topics raised by the Chair and Co-Chairs, relevant issues in CRC management were brought up. CN and LM developed a search strategy, and CN performed a systematic search on 4 databases and 11 organizations to source for CRC-related clinical practice guidelines to guide the development of our questionnaire items. The detailed search strategy used can be found in the **Supplementary Table A** below. 14 eligible guidelines were identified, and relevant information was extracted from these guidelines to evaluate the present state of research on CHM use in CRC management. The PRISMA flowchart depicting our search is shown in **Figure 2** below. CN reviewed relevant literature and drafted preliminary topics and clinical questions to be discussed at the following meetings. For the subsequent two meetings, the evidence was presented to the study team and based on further discussions, the draft consensus statements and questions were developed and shared with all members of the steering committee for review and refinement. Upon completion of the first Delphi survey draft, the questionnaire was translated by CN and LM onto an online survey format hosted on Microsoft Forms and piloted among four independent junior Chinese Medicine physicians not involved in our study. They were selected because of their familiarity with the Microsoft Forms platform used to host the survey and clinical familiarity with the topic. No changes were made to the questionnaire after review of technical issues, and their responses were not included in the final analysis.

**Figure 2:**
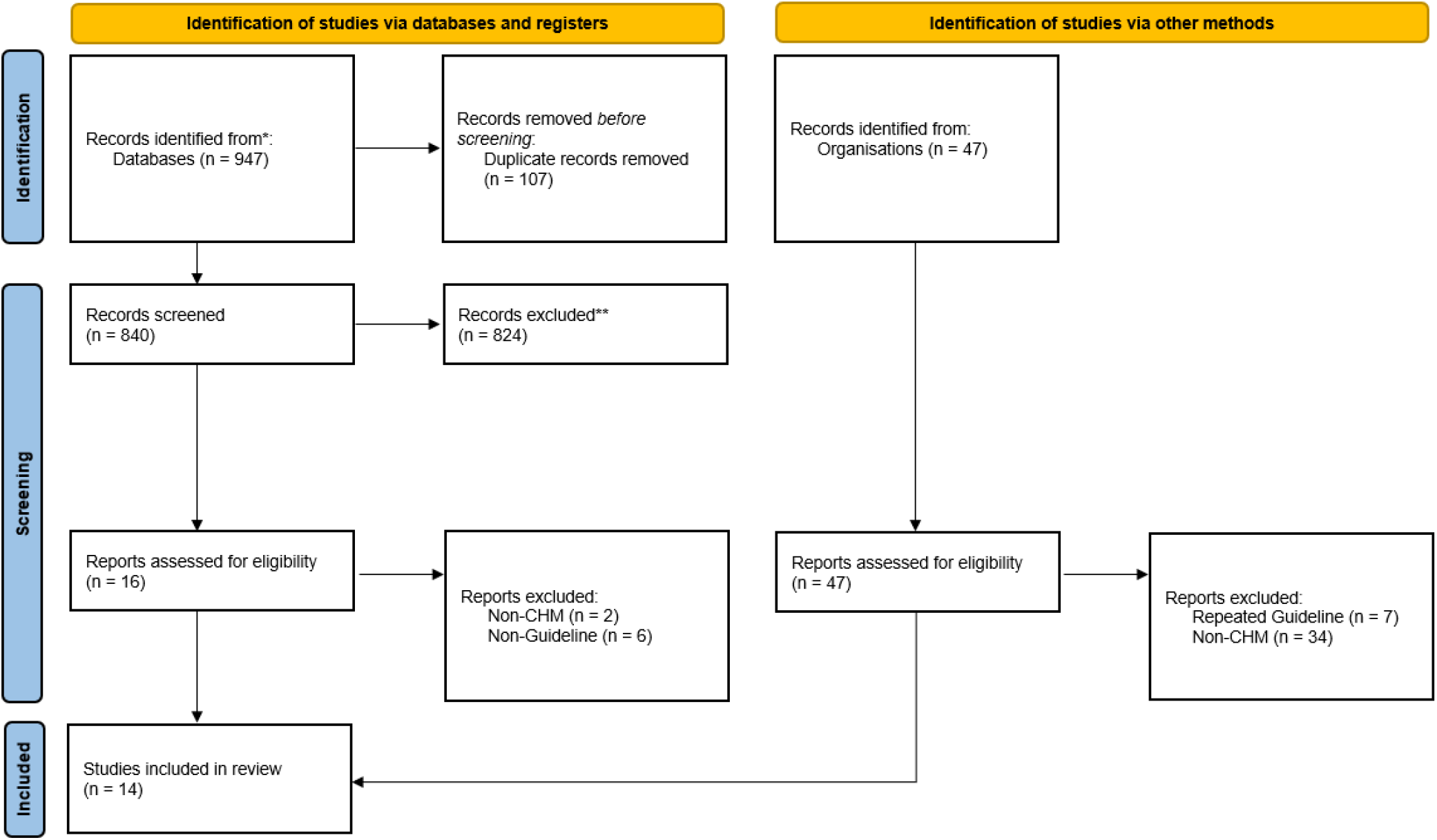
PRISMA flowchart depicting the results of our search

For the conduct of the Delphi consensus, a *“Survey Questions”* document and a *“List of Evidence”* document was prepared for the expert panelists. The “*List of Evidence*” document was prepared following the development of the questionnaire items, by manually searching the existing retrieved guidelines and their reference lists, and further on the Web of Science Core Collection database for high-quality evidence which supported each individual questionnaire item. The *“Survey Questions”* document consisted of two main classes of questions, namely “Opinion-type” questions and “Evidence-type” questions. The “opinion-type” questions required the expert panelists to answer the questions based on their clinical experience, while evidence questions required the expert panelists to refer to the *“List of Evidence”* document before providing their response. In addition, a free text box was also provided at the end of every “Evidence-type” question to allow panelists to provide feedback on the statements. A sample of the question construct is shown in **Textbox 2** below. The full survey questionnaire is attached as **Supplementary File 1**.

**[Textbox 2:** Constructs of sample questions]

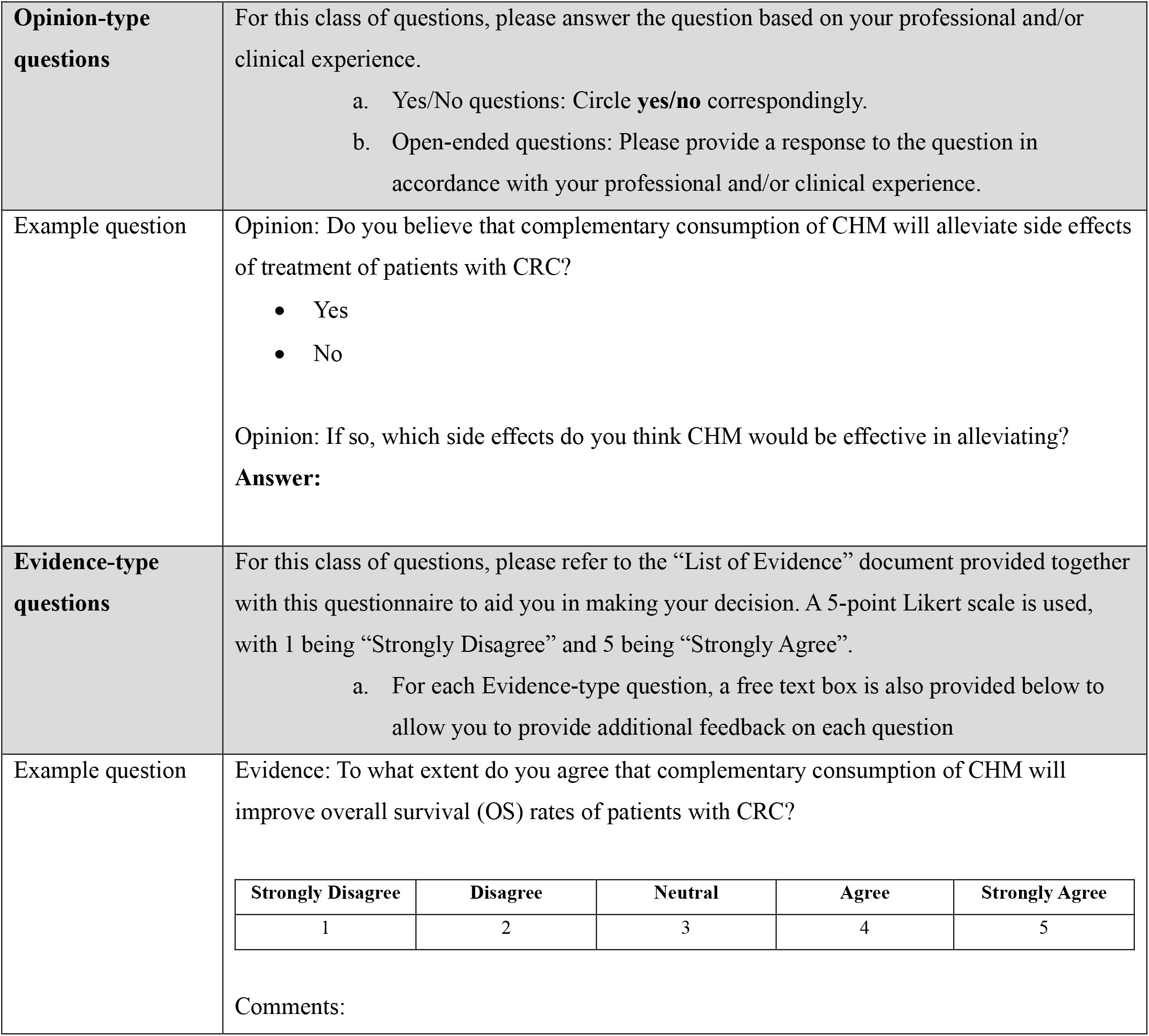

### Assessing consensus

The Delphi method was used to establish consensus. Our survey consisted of two main classes of questions, namely “Opinion-type” questions and “Evidence-type” questions. The “Opinion-type” questions required the expert panelists to answer the questions based on their clinical experience, while the “Evidence-type” questions required the expert panelists to refer to the *“List of Evidence”* document before providing their response.

Our study plans to use two rounds of Delphi survey to assess consensus. Level of agreement was rated on a scale from 1 (strongly disagree) to 5 (strongly agree), and free text fields allowed experts to provide additional feedback on each statement. Consensus was reached when ≥50% of experts surveyed voted with an agreement level of 4 or 5, and the level of consensus among experts was further graded using criteria in **Table 2**. For Round 1 of Delphi, the questions presented to our panelists are designed to be a mix of both statements and open-ended questions, allowing the panel to identify key issues and respond based on their experience and interpretation of the current evidence and practices. After the first round, statements which achieved “Strong Consensus” in the first round would be excluded from the second round. In the second round, the remaining statements will be presented to the panelists for a final consensus evaluation. For both rounds, the survey will be sent online via email for the panelists to fill up and return to the study team. Other than basic demographic information, the survey response will be anonymized, individually collected, and tabulated. Upon completion of Round 1, a written summary will be provided to the panel, and the feedback was aggregated across all expert groups. The results showing dissent were presented again for evaluation in the next Delphi round. Using answers from Round 1, Round 2 questions (statements only) will then be re-generated and sent to the expert panel again. The steering committee will not be involved in decisions made by the consensus panel throughout the whole Delphi process.

**Table 2:** Definitions for the level of agreement and the level of consensus for each statement.

| Level of agreement | Definition |
| --- | --- |
| 1 | Strongly disagree |
| 2 | Disagree |
| 3 | Neutral |
| 4 | Agree |
| 5 | Strongly agree |
| Level of consensus for each statement | Definition |
| Strong consensus | 76-90% of experts voted with agreement Level 4 or 5 |
| Consensus | 50-75% of experts voted with agreement Level 4 or 5 |
| No consensus | <50% of experts voted with agreement Level 4 or 5 |

### Participation

For this study, potential participants will be identified by the steering committee (LD, MX), and eligible experts will be invited to participate via an official email sent from the study team. The invitation email will include a brief introduction to the study objectives, an overview of the Delphi process, the anticipated time commitment, and information regarding confidentiality, voluntary participation, and the right to withdraw at any time without penalty. No remuneration will be provided for participation. All study-related communication will be conducted via email to ensure consistency, transparency, and traceability of correspondence. Participants who agree to take part will be required to provide informed consent electronically prior to commencement of the first Delphi round. To facilitate engagement, encourage responses and ensure a shared understanding of study procedures, a study e-guide was designed and sent to the survey participants about the conduct of the study. Thereafter, the consensus survey questions will be circulated online for the respondents to fill up for the next two Delphi rounds. To minimize response bias during the conduct of the survey and maintain the reliability of responses administered through the online questionnaire, “attention check” questions will be inserted at various intervals of the online survey [34]. Survey responses will be verified by CN at the end of each Delphi round to ensure consistency and reliability of the responses.

To maximise retention across Delphi rounds, reminder emails will be sent at two predefined intervals (3 days prior and 1 day prior to the deadline) to participants who have not yet completed each round’s questionnaire. Extension will be granted to participants who have not yet completed the responses on a case-by-case basis determined by the steering committee. These reminders will be limited to a maximum of two per round to minimise participant burden. No additional contact outside the agreed communication channels will be undertaken. Participants who do not respond after the final reminder will be considered lost to follow-up (LTFU) for that round, and their data will be excluded from subsequent rounds while retained in analyses of completed rounds where applicable. Non-compliance or partial responses will be handled by including only complete responses in each round’s analysis.

### Sample size determination

As Delphi studies do not rely on statistical power calculations, panel size will be determined based on methodological guidance and anticipated attrition across rounds. Based on previous recommendations, the panel size should consist of a minimum of 20 members [26-29]. Hence, we aim to have 25–30 experts completing the final Delphi round. Allowing for ∼20–30% attrition, we plan to recruit approximately 35–50 experts by invitation.

### Statistical plan

All questionnaire data will be exported from the online survey platform into a secured database for cleaning and analysis. Responses will be analysed by each Delphi round. Only completed responses for each round will be included in that round’s analysis. Participants who do not complete a round after the predefined reminder schedule will be classified as LTFU for subsequent rounds. In the analysis, the mean values for percent agreement are weighted for each expert group in terms of the number of group members. However, their completed prior-round responses will be retained for analysis of those rounds. The software used for statistical analysis will be R. A breakdown of the different statistical analyses for each juncture of the study is shown in **Table 3** below.

**Table 3:** Statistical analysis procedure at each study juncture.

| Study item | Analysis procedure |
| --- | --- |
| Panellist characteristics | Will be summarised using counts and percentages for categorical variables and median (IQR) for continuous variables. |
| Opinion-type questions | For items with dichotomous responses (Yes/No), responses will be summarised at the statement level using counts and percentages of “Yes” and “No”, together with the total number of respondents (n) for that round. |
| Evidence-type questions | Item responses will be summarised using:<br>• n (%) in each response category (Levels 1–5)<br>• Median (IQR) of the Likert score<br>• Agreement proportion, defined as the percentage of respondents selecting Level 4 or Level 5 of agreement |
| Free-text responses | Will be analysed using content analysis. Comments will be coded into themes that inform: (i) refinement of existing statements, (ii) generation of new statements (if applicable), and (iii) clarification of definitions or implementation considerations. The number of comments contributing to revised newly generated statements will be recorded. |
| Attrition and response stability | Response rates will be reported for each Delphi round, including overall attrition across rounds. To describe response stability, we will report round-to-round changes in:<br>• The agreement proportion (Level 4/5), and<br>• The consensus category assigned to each statement (e.g., no consensus/consensus/strong/) |

## Discussion

Through the conduct of this Delphi study, we aim to establish Expert Consensus on key clinical concerns guiding the usage of CHM as a complementary therapy in the management of CRC. We anticipate that establishing consensus on key clinical concerns such as determining the (1) optimal intervention, (2) recommended stage and duration of intervention, (3) safety considerations, and (4) possible herb-drug interactions, will serve to lay important groundwork for potential future incorporation of CHM into CRC treatment protocols alongside conventional oncology approaches. Following the successful completion of the study, we also anticipate the conversion of the results from the Expert Consensus into a clinical pathway or clinical practice guideline for complementary CHM in CRC management.

### Dissemination Plan

The findings of this study will be disseminated through peer-reviewed publications and conference presentations.

### Delphi Study Status

The literature review was completed in November 2025, together with the development and finalization of the survey items in December 2025. This study has obtained IRB approval from Nanyang Technological University before commencement (IRB-2025-1222).

## Data Availability

All data produced in the present study are available upon reasonable request to the authors

## Ethics Approval

Institutional Review Board, Nanyang Technological University (Application: IRB-2025-1222). Informed consent will be obtained for each participant at the beginning of the survey, and participants will be free to withdraw from the study at any point of the survey without any consequences.

## Author contributions

**CYJN:** Conceptualization, Methodology, Visualization, Writing - Original Draft, Writing - Review & Editing.

**ML:** Methodology, Writing - Review & Editing.

**DA:** Writing - Review & Editing.

**YL, MY**: Methodology, Resources, Writing - Review & Editing, Supervision.

**LLDZ:** Conceptualization, Methodology, Resources, Writing - Review & Editing, Supervision. All authors have read and approved this manuscript for publication.

## Supplementary Information

**Supplementary Table A:**
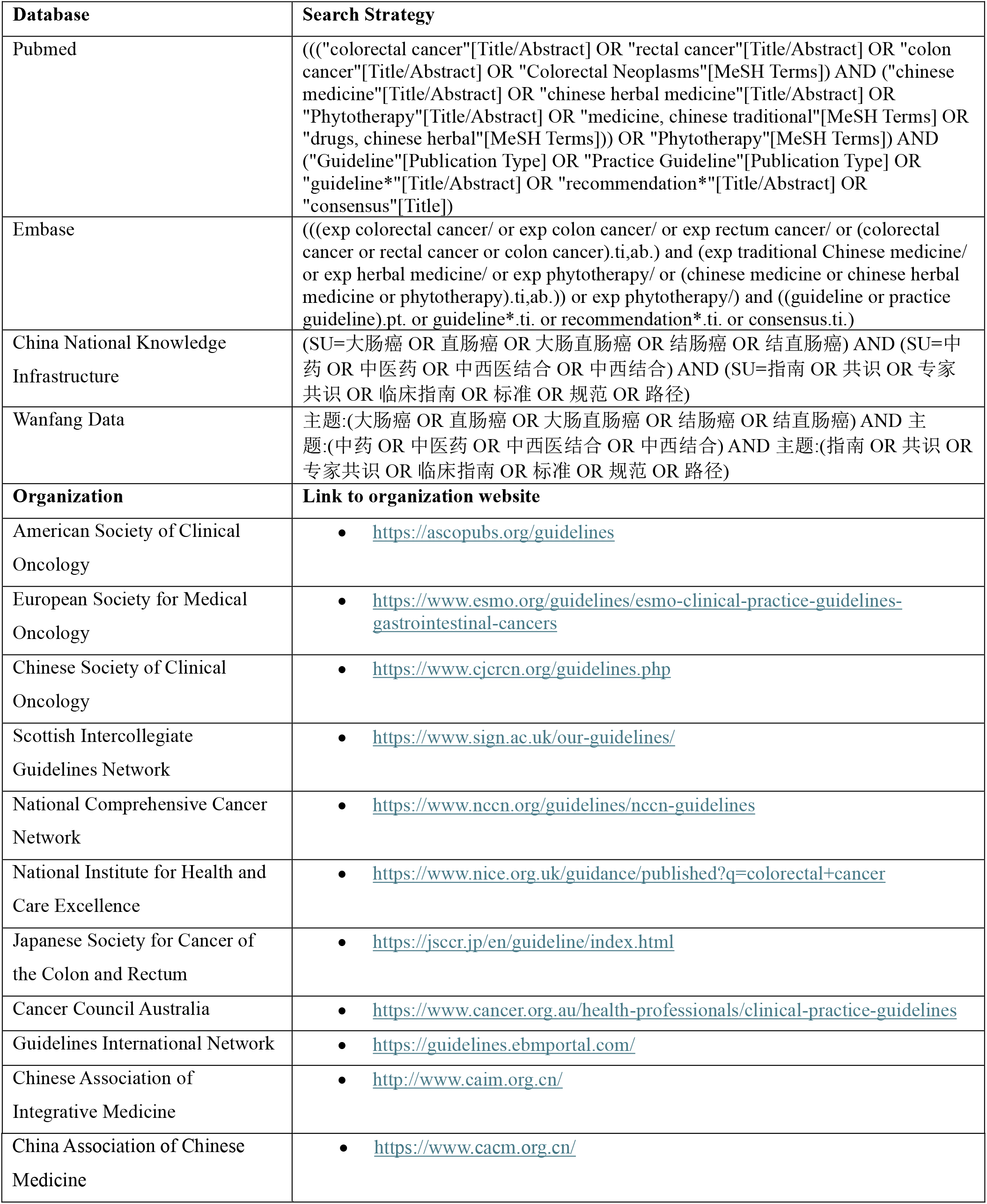
Detailed search strategy of databases and organizations.

